# Can GPT Be Used as an Alternative to Traditional Machine Learning and Neural Networks for Prediction on Small Clinical Datasets?

**DOI:** 10.64898/2026.08.19.26360765

**Authors:** Simon Bin Akter, Sumya Akter, David Eisenberg, Chelsey Hill, Aaron Lotvola, Jorge Fresneda Fernandez, Tanmoy Sarkar Pias, Muhammad Rafiqul Islam, Humayera Islam

## Abstract

**Purpose:** Early and reliable disease prediction from structured clinical data remains challenging when datasets are small, highly imbalanced, and contain limited positive disease cases. Conventional machine learning (ML) and deep learning approaches often struggle to capture clinically meaningful relationships under such low-data representation conditions due to weak statistical associations between features and prediction targets. This study proposes a clinically grounded Distil GPT2-based table-to-text framework for disease prediction using structured healthcare datasets, motivated by the contextual reasoning capability of GPT models to better capture clinically meaningful relationships when statistical learning alone becomes insufficient due to limited data availability.

**Methods:** Structured clinical records were transformed into physician-style textual descriptions and enriched through GPT4-generated medical paraphrasing to improve minority-class representation while preserving clinical meaning. Both the original and generated clinical texts were used to fine-tune a Distil GPT2 model across four public healthcare datasets, including heart disease, heart failure, chronic kidney disease, and thyroid cancer recurrence. Gradient-based explainable AI analysis was additionally incorporated to identify clinically important features influencing prediction outcomes.

**Results:** The proposed framework demonstrated consistently strong predictive performance across four clinical datasets, achieving average precision, specificity, sensitivity, and F1-score of 0.96, 0.97, 0.96, and 0.96, respectively. The model achieved improved sensitivity, stronger generalization, and more stable predictive behavior compared with traditional ML, deep learning, transformer-based, and GAN-augmented approaches. Importantly, the framework consistently emphasized clinically meaningful variables even under severe class imbalance, where conventional ML and neural network models often struggled to identify key clinically relevant relationships.

**Conclusion:** The proposed Distil GPT2-based tableto-text framework provides a practical and clinically interpretable approach for disease prediction from limited structured healthcare data. By integrating contextual clinical reasoning with explainable prediction mechanisms, the framework suggests strong potential for early risk detection, transparent clinical decision support, and reliable deployment in real-world data-scarce healthcare settings.

## 1 INTRODUCTION

Health conditions such as heart disease, heart failure, kidney disease, and differentiated thyroid cancer recurrence are rapidly increasing worldwide, making prevention strategies and largescale early detection essential for improving public health outcomes [1]. Machine learning (ML)–based approaches can be highly effective in enabling cost-effective and widespread disease screening [2]. In the modern era, the increasing volume of structured clinical datasets has created immense potential for data-driven decisionmaking in healthcare [3]. These datasets can be highly beneficial for ML-based disease prediction tasks, enabling early, accurate, and affordable detection to reduce mortality and improve treatment outcomes [4]. However, a major challenge lies in effectively utilizing these tabular records, which are typically composed of categorical and numerical values and often suffer from class imbalance and underrepresentation of disease cases [2]. ML-based prediction models often struggle to produce reliable results when disease cases are underrepresented, as they rely solely on statistical relationships between categories and cannot capture the subjective clinical context [5]. In scenarios where datasets exhibit very low representation of disease samples [6, 7], subjective, context-aware modeling, such as large language models (LLMs) [4, 8], can be more beneficial than purely statistical ML or neural network–based methods. LLMs can ensure a better understanding of the data by capturing both the statistical patterns and the subjective clinical context [9].

Recent advances in healthcare analytics have largely relied on classical ML and ensemble models applied to structured clinical data [3, 4]. While these approaches have demonstrated strong predictive performance, particularly through recall optimization under class imbalance, their evaluation remains predominantly metric-driven [10]. Clinical variables are typically treated as independent numerical or categorical features, and model success is measured almost exclusively through statistical indicators such as accuracy, recall, and F1-score [2]. However, this performance-centric paradigm raises critical concerns regarding model trustworthiness and clinical reliability [11]. Aggressive recall optimization, often achieved via resampling or cost-sensitive learning [5], can conceal subtle but clinically meaningful interactions among symptoms, comorbidities, and laboratory findings. Moreover, most existing studies lack mechanisms for contextual reasoning or clinically grounded explanations, limiting their ability to support transparent and trustworthy medical decision-making [12–14]. This gap highlights a fundamental limitation of correlation-driven learning in healthcare: high predictive scores do not necessarily translate into clinically interpretable or reliable risk assessment [5, 11]. Addressing this challenge requires modeling frameworks that move beyond tabular pattern fitting toward context-aware, explainable representations of clinical data, enabling both robust sensitivity and meaningful clinical insight [11].

In this context, the emergence of pretrained LLMs, particularly GPT-based architectures [15], presents a promising alternative for clinical prediction tasks involving limited or low-representation datasets. Unlike traditional ML and neural network models [8, 16] that rely heavily on sufficient positive-case distribution to learn stable feature associations, pretrained LLMs leverage extensive prior knowledge acquired from large-scale biomedical and general-domain corpora. This broad pretrained knowledge enables GPT models to capture contextual clinical relationships alongside statistical feature interactions, allowing them to reason more effectively about symptom combinations, disease progression patterns, laboratory abnormalities, and comorbid conditions even under data-scarce settings [17]. Consequently, GPT-based models can provide more clinically reliable interpretations of feature relevance and disease risk compared with purely correlation-driven learning methods. Their ability to integrate contextual understanding with predictive reasoning offers a potential pathway toward improving both explainability and clinical trustworthiness in low-volume healthcare datasets, where conventional ML and deep neural networks often struggle to generalize reliably due to insufficient representation of positive clinical cases [6].

This study investigated whether GPT can make more reliable disease predictions from small and imbalanced clinical datasets [6] by learning medical context rather than only statistical patterns. Instead of using raw tables, each patient’s clinical record was converted into short, physician-style text descriptions that preserved the original medical meaning. These texts were then carefully paraphrased using a GPT-based model to generate realistic new patient examples, improving data balance without altering clinical facts. A Distil GPT2 model was subsequently fine-tuned on these clinical texts and evaluated across multiple public healthcare datasets. Importantly, to make the model’s decisions more transparent, a gradient-based explanation method was used to identify which clinical terms most influenced each prediction, highlighting key medical features that the model relied on, so the output supports not only prediction but also clinically understandable reasoning. The model demonstrated improved sensitivity, stronger generalization, and clearer use of meaningful clinical features compared to traditional ML methods and neural networks. By combining statistical learning with clinically grounded language understanding, this approach offers a more trustworthy and interpretable framework for disease prediction from limited clinical data.

The key contributions are summarized below:

1. **Clinical narrative representation for tabular prediction:** Converts numerical and categorical patient records into physician-style descriptions to leverage Distil GPT-2’s context-aware language representations and address learning challenges associated with limited minority-class representation.
2. **Fact-preserving minority-class augmentation:** Uses GPT-4 paraphrasing to introduce linguistic diversity while preserving clinical measurements, units, and class labels.
3. **Joint evaluation of predictive performance and clinical feature relevance:** Assesses predictive metrics and clinically relevant token attributions alongside feature explanations from strong conventional baselines.
4. **Evaluation across four clinical datasets with medical expert assessment:** Compares the framework with machine learning, neural networks, alternative language models, and tabular augmentation methods across four clinical prediction tasks. Medical experts additionally assess the clinical relevance of the identified features, supporting interpretation of the study’s findings.

By transforming structured clinical records into clinically meaningful text and integrating transparent explanation mechanisms, this work moves beyond black-box prediction toward reliable clinical decision support. The proposed approach enables healthcare professionals to not only identify high-risk patients early, even in data-scarce settings, but also understand why a prediction was made through clinically relevant indicators. This transparency strengthens trust, supports informed clinical judgment, and reduces reliance on purely statistical correlations. As a result, the framework offers a practical pathway for deploying language-model–based tools in realworld healthcare systems, where interpretability, reliability, and clinical accountability are essential for safe and effective adoption, summarized in **Table 1**.

**Table 1:** Statement of significance.

|  |
| --- |
| **Problem or Issue:**<br>Disease prediction from small and imbalanced clinical datasets often produces unreliable and poorly interpretable results. |
| **What is Already Known:**<br>Traditional machine learning and deep learning models primarily learn statistical correlations, which struggle to preserve clinically meaningful relationships when positive cases are limited. |
| **What this Paper Adds:**<br>This study introduces a Distil GPT-2 table-to-text framework that combines statistical learning with context-aware language understanding to capture clinically meaningful patterns beyond numerical correlations. Clinical paraphrasing improves minority-class learning, while gradient-based explanations provide transparent reasoning, supporting more reliable and interpretable clinical decision-making. |

The remainder of this paper is organized as follows. The literature review section summarizes related work on ML and language model–based approaches for disease prediction from clinical data. The materials and methods section describes the proposed framework, including dataset descriptions, data preprocessing, clinical text generation, model training, and the explanation strategy. The results and analysis section presents the experimental findings, covering both predictive performance and interpretability analysis [18]. The discussion section highlights the clinical implications, strengths, and practical significance of the proposed approach. Finally, the conclusion section summarizes the major findings of the study and their potential impact on real-world healthcare applications.

## 2 LITERATURE REVIEW

Studies using the heart disease dataset primarily apply classical ML and ensemble models to structured clinical attributes [19–21]. The focus is largely on optimizing performance metrics, particularly accuracy, recall, and F1-score, often through feature selection, resampling techniques, or model ensembling. These approaches treat clinical variables as independent numerical or categorical inputs and evaluate success mainly through statistical performance.

Research on heart failure datasets typically emphasizes predicting mortality or disease progression using tabular clinical data and supervised ML classifiers [22–24]. Many studies prioritize recall and sensitivity due to the high clinical cost of false negatives, frequently addressing imbalance using oversampling or cost-sensitive learning. However, the models remain correlation-driven, with limited insight into how combinations of symptoms, comorbidities, and lab measurements contribute to clinical risk.

Chronic kidney disease prediction studies rely heavily on traditional ML pipelines involving preprocessing, imputation, feature selection, and classifiers [25–28]. Performance improvements are often achieved by balancing datasets and tuning hyperparameters. While high recall is reported, these models generally lack mechanisms to capture longitudinal disease context or clinically meaningful interactions among laboratory and demographic features.

Existing work on differentiated thyroid cancer recurrence prediction applies ML and ensemble methods to clinicopathologic tabular datasets, frequently using traditional sampling techniques to mitigate class imbalance [29, 30]. These studies report very high accuracy and recall. However, the modeling remains dataset-specific and statistically driven, with interpretability limited to feature importance scores and no explicit clinical reasoning or explanation of recurrence risk.

For Heart Disease, the highest reported precision is 95.65%, sensitivity is 91.70%, specificity is 97.22%, and F1-score is 93.61% [19–21]. For Heart Failure, the highest reported precision is 91%, recall is 93%, and F1-score is 90% [22–24]. For Chronic Kidney Disease, the highest reported precision is 99.40%, recall is 99.41%, and F1-score is 99.61% [25–28]. For Recurrence in Differentiated Thyroid Cancer, the highest reported accuracy is 98.17% [29, 30]. Across these datasets, existing studies [19–30] are primarily focused on improving recall under class imbalance, often equating this with better clinical learning. However, our analysis reveals that aggressively optimizing recall, often via oversampling, can cause models to overlook important but subtle clinical patterns and interactions present in the data. Moreover, current approaches provide little to no clinical explainability, relying entirely on statistical associations without modeling subjective medical context or reasoning. To address this gap, we adopt a tableto-text context-aware modeling paradigm that goes beyond recall maximization by enabling the model to learn and explain clinically meaningful patterns from imbalanced tabular data. This allows improved sensitivity without sacrificing the learning of critical clinical relationships, while also providing interpretable, clinically grounded decision support rather than purely correlation-based predictions. Recent table-to-text studies primarily focus on generating coherent natural language from structured tables using hierarchical content planning, with evaluations largely conducted in domains such as sports analytics to ensure factual fidelity and narrative coherence [31, 32]. However, these approaches are largely developed under the premise of balanced and complete tables and do not address skewed data distributions or the need to reliably capture rare but clinically critical patterns, as addressed in this study.

## 3 MATERIALS AND METHODS

This methodology section describes each step of the proposed study, presented in **Fig 1**. It first presents the clinical datasets, feature representations, and train-test formulation used in the study. Next, table-to-text clinical transformation and GPT-4–based synthetic text generation were applied to address class imbalance while preserving medical reliability. A context-aware Distil GPT2 framework was then developed for clinical prediction and compared with multiple ML, deep learning, transformer, and GAN-based approaches. Finally, interpretability analysis was performed using gradient-based token attribution and SHAP-based feature importance to evaluate contextual clinical reasoning across different models. For additional examples of clinical text conversion from tabular data and synthetic positive-case generation through paraphrasing, please refer to **Fig. A.1, Fig. A.2, Fig. A.3**, and **Fig. A.4**, which present examples from the four datasets, respectively, in the Supplementary section.

**Fig. 1:**
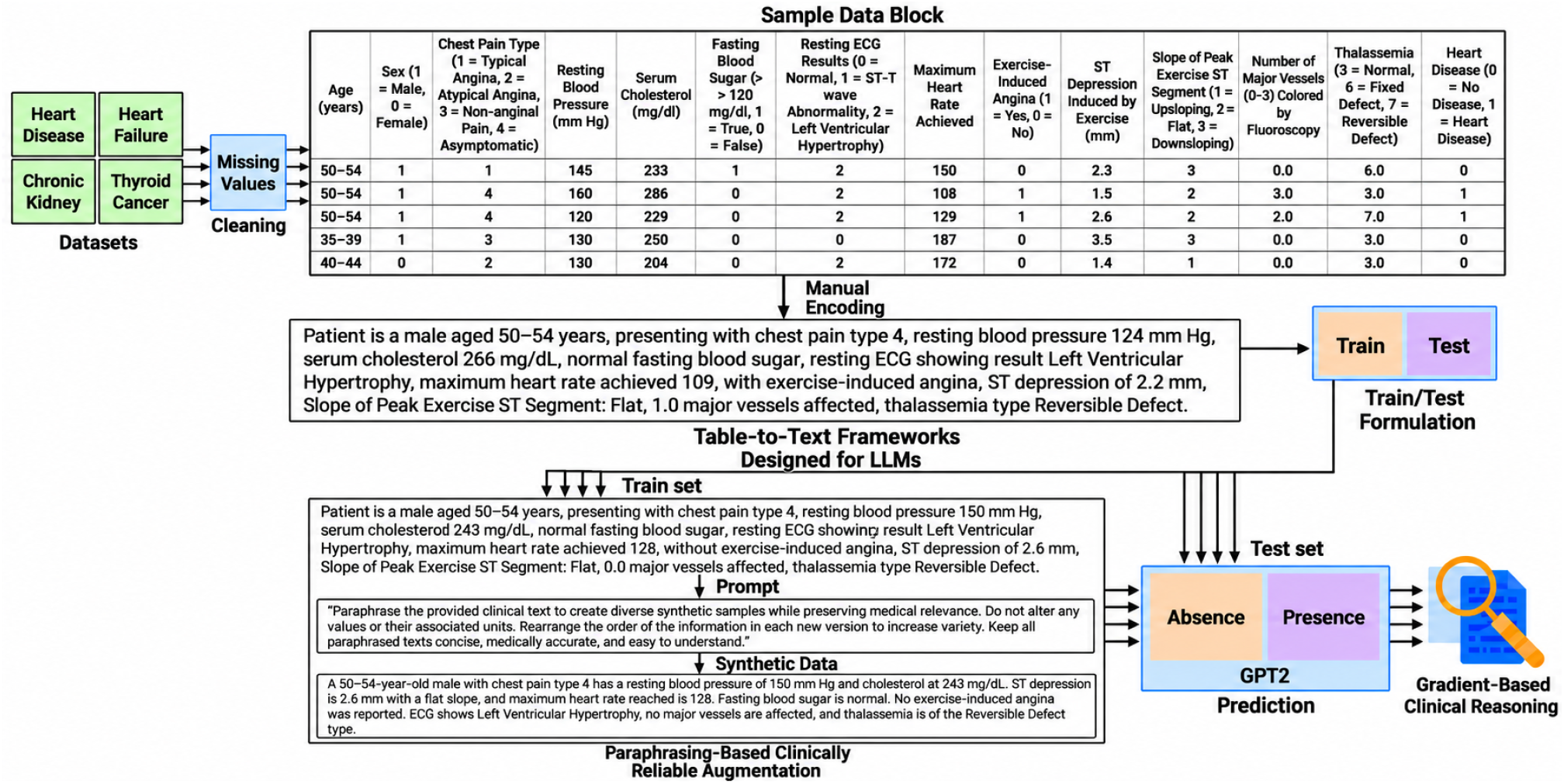
Complete workflow of the proposed Distil GPT2-based table-to-text clinical prediction framework. Structured clinical records are transformed into physician-style textual descriptions and augmented through GPT4-based clinical paraphrasing to improve minority-class representation under severe class imbalance. The generated clinical text is then used to fine-tune a Distil GPT2 model for disease prediction across multiple healthcare datasets. Finally, gradient-based explainable AI analysis is employed to identify clinically important features and provide transparent, context-aware reasoning for reliable medical decision support.

### 3.1 Data Description

This study utilizes four publicly available datasets from the UCI Machine Learning Repository to evaluate model performance and reasoning across multiple clinical conditions. The UCI Heart Disease dataset [33] contains 303 samples, where the target variable represents the presence (1) or absence (0) of heart disease, comprising approximately 139 positive and 164 negative cases. The UCI Heart Failure Clinical Records dataset [34] includes 299 samples, where the target variable indicates mortality outcome, consisting of 96 deceased cases and 203 surviving cases. The UCI Chronic Kidney Disease dataset [35] consists of 400 samples, with the target variable representing disease presence versus absence, including 250 disease-positive and 150 disease-negative cases. Finally, the UCI Differentiated Thyroid Cancer Recurrence dataset [36] comprises 383 samples, where the target variable denotes recurrence status, including approximately 108 recurrence cases and 275 non-recurrence cases.

### 3.2 Feature Description

In **Table 2**, summarizes the major clinical features and target variables of the four datasets [33–36] used in this study. The datasets include demographic information, physiological measurements, laboratory biomarkers, disease history, and clinical examination findings relevant to their respective prediction tasks.

**Table 2:** Clinical datasets and corresponding feature descriptions used in this study.

| Dataset | Main Features | Target Variable |
| --- | --- | --- |
| Heart Disease | Age, sex, chest pain type, resting blood pressure, cholesterol level, fasting blood sugar status, resting electrocardiographic results, maximum heart rate achieved, exercise-induced angina, ST depression, slope of ST segment, number of major vessels observed through fluoroscopy, and thalassemia-related measurements. | Presence or absence of heart disease |
| Heart Failure Clinical Records | Age, anemia status, creatinine phosphokinase level, diabetes status, ejection fraction, hypertension status, platelet count, serum creatinine level, serum sodium level, sex, smoking status, and follow-up duration. | Mortality outcome during follow-up |
| Chronic Kidney Disease | Age, blood pressure, specific gravity, albumin level, sugar level, red blood cells, pus cells, bacteria presence, blood glucose, blood urea, serum creatinine, sodium, potassium, hemoglobin, packed cell volume, white blood cell count, red blood cell count, hypertension, diabetes mellitus, coronary artery disease, appetite condition, pedal edema, and anemia. | Presence or absence of chronic kidney disease |
| Differentiated Thyroid Cancer Recurrence | Age, gender, smoking history, radiation therapy history, thyroid function, physical examination findings, adenopathy condition, pathology type, tumor focality, risk category, tumor staging parameters, overall cancer stage, and treatment response. | Occurrence of thyroid cancer recurrence |

### 3.3 Table-to-Text and Train/Test Formulation

After cleaning the missing values, each tabular record in the dataset was transformed into a clinical-style textual representation on a row-wise basis to make it compatible with language models. For each row, all clinical variables, their associated values, and labels are formatted descriptively as structured patient history notes without altering or modifying the original data. These text representations were manually constructed and subsequently verified by medical experts and clinicians to ensure clinical correctness and consistency represented in **Fig 1**. Further examples of converting tabular clinical data into contextual clinical text are presented in **Fig. A.1, Fig. A.2, Fig. A.3**, and **Fig. A.4** in the Supplementary section, corresponding to the four datasets, respectively. Then, the datasets were randomly split into an average ratio of 82:18 for training and testing, respectively, while carefully ensuring that no data leakage occurred between the two sets. This ratio was selected to maintain a sufficient number of positive-class samples in both the training and testing sets, as the availability of positive-class data was highly limited. For the different train–test configurations, please refer to **Table B.1** in the Supplementary section.

### 3.4 Clinical Text Generation

The training sets exhibited substantial class imbalance, particularly for the positive and clinically important classes, presented in **Table 3**. To address this issue without applying conventional resampling techniques such as random oversampling, undersampling, or interpolation methods that can affect medical reliability, a web-based GPT-4 model [3, 4, 8, 16] was used to generate diverse synthetic textual samples through controlled paraphrasing. Each clinical text record was individually provided to the model using the following prompt: “Paraphrase the provided clinical text to create diverse synthetic samples while preserving medical relevance. Do not alter any values or their associated units. Rearrange the order of the information in each new version to increase variety. Keep all paraphrased texts concise, medically accurate, and easy to understand.”

**Table 3:** Class distributions in the training and test sets before and after training-set augmentation. These counts reflect the data remaining after removing records with missing values. Augmentation was applied only to the training data; test sets retained their original class distributions. The number of augmented samples was determined through iterative experiments.

| Dataset | Training set<br>before augmentation |  | Training set<br>after augmentation |  | Test set<br>unchanged |
| --- | --- | --- | --- | --- | --- |
|  | Negative | Positive | Negative | Positive | Negative \|<br>Positive |
| Heart Disease | 128 | 118 | 128 | 164 | 32 \| 19 |
| Heart Failure<br>Clinical Records | 162 | 87 | 162 | 172 | 41 \| 9 |
| Chronic Kidney<br>Disease | 92 | 34 | 92 | 92 | 23 \| 9 |
| Differentiated<br>Thyroid Cancer<br>Recurrence | 220 | 91 | 220 | 220 | 55 \| 17 |

The generated synthetic samples modified only the linguistic phrasing and sentence structure while preserving all original clinical variables, numerical values, units, and class labels represented in **Fig 1**. Further examples of clinical text generation are presented in **Fig. A.1, Fig. A.2, Fig. A.3**, and **Fig. A.4** in the Supplementary section, corresponding to the four datasets, respectively. Furthermore, all generated synthetic texts were subsequently reviewed and verified by medical experts and clinicians to ensure medical accuracy, consistency, and preservation of the original clinical meaning. This approach aimed to increase the representation of minority and clinically important classes without altering the underlying medical information, thereby maintaining clinical reliability and avoiding the risks associated with traditional synthetic data generation techniques.

### 3.5 Context-Aware Modeling

In this study, a context-aware clinical prediction framework was developed using the Distil GPT2 architecture obtained from the Hugging Face Transformers library. The model was trained on clinical-style textual representations generated from tabular medical records along with their corresponding synthetic paraphrased samples. The framework was evaluated across four clinical prediction tasks, including heart disease detection, heart failure survival prediction, chronic kidney disease identification, and differentiated thyroid cancer recurrence prediction.

The motivation behind using a transformer-based language model instead of relying solely on traditional ML and neural network approaches originates from the severe class imbalance and low representation of clinically important cases within the datasets [5, 6]. Conventional ML algorithms and standard neural networks primarily depend on learning statistical relationships directly from the available training samples [2]. However, when minority-class samples are extremely limited, the statistical relevance between features and target labels becomes weak and unstable, often reducing generalization capability [5, 6].

In contrast, Distil GPT2 is pretrained on massive textual corpora and therefore possesses extensive contextual and semantic knowledge learned during pretraining [37]. This prior knowledge enables the model to capture contextual clinical relevance in addition to purely statistical feature correlations. Consequently, even when clinically important cases have very limited representation, Distil GPT2 can still identify meaningful clinical patterns and dependencies that can influence prediction outcomes. This context-aware capability reduces the likelihood of missing important information due to insufficient data representation and helps maintain more stable predictive behavior in low-resource clinical settings.

To adapt Distil GPT2 for supervised clinical prediction tasks, the original autoregressive language modeling head was replaced with a classification layer. The input clinical text sequence can be represented as:

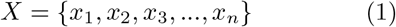

where *x*_*i*_ denotes the *i*^*th*^ token in the clinical text sequence.

The input sequence is processed through stacked masked self-attention transformer layers:

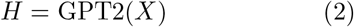

where *H* represents the contextual hidden representations generated by Distil GPT2:

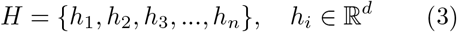

The hidden representation of the final token was used as the aggregated contextual representation of the entire clinical sequence:

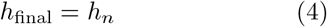

A dropout layer was then applied for regularization:

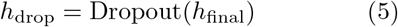

The contextual representation was projected into the output prediction space using a linear classification layer:

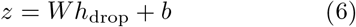

where *W* and *b* represent the learnable classifier weight matrix and bias vector, respectively.

The probability distribution over the prediction classes was computed using the softmax activation function:

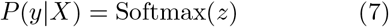

The model parameters were optimized using the cross-entropy loss function:

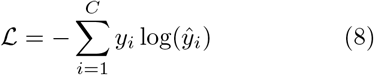

where *C* denotes the number of classes, *y*_*i*_ represents the ground-truth label, and ŷ_*i*_ denotes the predicted probability for class *i*.

Internally, Distil GPT2 utilizes masked multihead self-attention to model contextual dependencies among clinical variables. The attention operation is defined as:

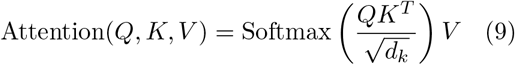

where *Q, K*, and *V* denote the query, key, and value matrices, respectively, and *d*_*k*_ represents the dimensionality of the key vectors.

To provide interpretability and transparency, a gradient-based post-hoc explanation approach was employed after model training [38]. The trained Distil GPT2 model was kept fixed, and the gradient of the predicted class score with respect to each input token embedding was computed as:

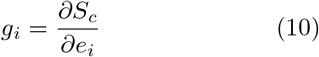

where *S*_*c*_ represents the predicted score for class *c*, and *e*_*i*_ denotes the embedding vector corresponding to token *i*.

The magnitude of the gradient, |*g*_*i*_| indicates how sensitive the prediction is to a specific clinical token. Larger gradient magnitudes imply that the model relied more heavily on that token during decision making. By focusing specifically on clinically meaningful keywords such as blood pressure, glucose, creatinine, cholesterol, ejection fraction, and related medical attributes, the framework highlights which clinical variables most strongly influenced the prediction outcome. This approach provides transparent and clinically interpretable explanations without modifying the original model architecture.

The reasoning patterns and interpretability behavior generated by Distil GPT2 were further compared with the interpretability results of the highest-performing traditional ML and neural network models across all four datasets. This comparison was conducted to investigate whether the contextual understanding capability of Distil GPT2 enables stronger identification of clinically relevant patterns under limited-data conditions compared with purely statistical learning approaches.

The pretrained DistilGPT-2 model was fine-tuned using a maximum sequence length of 128 tokens, a per-device batch size of 2, gradient accumulation of 2, weight decay of 0.01, a dropout rate of 0.2, and a warm-up ratio of 0.1. FP16 mixed-precision training [39] was enabled to reduce computational and memory requirements. Training comprised 730, 664, 460, and 660 optimization steps for the UCI Heart Disease, Heart Failure Clinical Records, Chronic Kidney Disease, and Differentiated Thyroid Cancer Recurrence datasets, respectively, corresponding to 10, approximately 7.91, 10, and 6 completed epochs. Fine-tuning was performed on an NVIDIA Tesla T4 GPU, with training times of approximately 23.1, 57.1, 8.0, and 18.0 minutes, respectively. The overall mean training losses were 0.7317, 0.6861, 0.3480, and 0.3801, respectively. The remaining hyperparameter settings were kept consistent across datasets.

## 4 RESULTS AND ANALYSES

The experimental evaluation first included multiple machine learning and deep learning approaches, including Adaptive Boosting (AdaBoost), Random Forest (RF), Decision Tree (DT), Gaussian Naive Bayes (GNB), Random Under Sampling Boosting (RUSBoost), Artificial Neural Network (ANN), Multi-Layer Perceptron (MLP), Convolutional Neural Network (CNN), transformer-based input sequence modeling, T5-small, FLAN-T5-Small, BART-Base, and Distil GPT2-based table-to-text modeling. Next, Synthetic Minority Over-sampling Technique for Nominal and Continuous Features (SMOTE-NC), Conditional Tabular Generative Adversarial Network (CTGAN), Tabular Variational Autoencoder (TVAE), and Copula Generative Adversarial Network (CopulaGAN) were applied for data balancing and augmentation [10]. Subsequently, model performance was evaluated using macro-averaging precision, specificity, sensitivity, and macro-averaging F1-score with 95% confidence intervals computed using a non-parametric bootstrap procedure with 1,000 resampling iterations. Finally, interpretability analysis was conducted using gradient-based token attribution for Distil GPT2 and Shapley Additive Explanations (SHAP) for the highest-performing ML and neural network models to compare contextual clinical reasoning and feature relevance identification.

### 4.1 Experimental Results: Performance

**Table 4** and **Table 5** present the comparative performance of traditional machine learning, deep learning, transformer-based, and GPT-based approaches across the four clinical prediction tasks under multiple synthetic data generation strategies. Overall, the Distil GPT2 (Table to Text) framework combined with GPT-4–generated clinical paraphrased samples demonstrated the most stable and consistently high performance across all datasets. In the Heart Disease dataset, the proposed approach achieved the highest overall F1-score while maintaining strong precision, specificity, and sensitivity. For the Heart Failure dataset, Distil GPT2 (Table to Text) with GPT4-generated samples substantially improved performance compared with conventional ML, ANN, CNN, and transformer input-sequence models, achieving the best overall balance between precision, specificity, sensitivity, and F1-score. In the Chronic Kidney Disease dataset, the proposed framework achieved perfect classification performance across all evaluation metrics, outperforming all Generative Adversarial Network (GAN)-based and resampling-based approaches. This is because the available testing sample size was small, and existing literature has also reported similar performance levels exceeding 99% [25–28]. Similarly, in the Thyroid Cancer Recurrence dataset, Distil GPT2 (Table to Text) with GPT4 augmentation achieved the highest precision and F1-score while maintaining perfect specificity. Additionally, across all four datasets, Distil GPT2 was compared with similarly sized language models, including T5-Small (60.51M parameters), FLAN-T5-Small (80.98M parameters), and BART-Base (139.42M parameters) [40]. When using the table-to-text formulation and comparable training epochs, Distil GPT2, with 82M parameters [41], consistently outperformed these comparable LLMs with similar training epochs. Other LLMs often performed comparatively better, but their performance was generally poor and highly unstable compared to Distil GPT-2.

**Table 4:** Performance comparison of different machine learning, deep learning, and GPT-based approaches across the Heart Disease and Heart Failure datasets under different synthetic data generation strategies. The best-performing models using sampling techniques are reported here. The 95% confidence interval is reported for each evaluation metric. Here, ± indicates the range within which the model’s performance can vary but it does not necessarily reach these limits, especially if its performance is already at or near a boundary. The ablation study also suggests how each individual component contributes to the overall performance of the proposed framework.

| Dataset | Model | GAN | Precision | Specificity | Sensitivity | F1 Score |
| --- | --- | --- | --- | --- | --- | --- |
| Heart Disease | FLAN-T5-Small (Table to Text) | GPT4 | 0.31 ± 0.11 | **1.00 ± 0.00** | 0.00 ± 0.00 | 0.39 ± 0.11 |
| | ANN | SMOTE-NC | 0.52 ± 0.14 | 0.41 ± 0.17 | 0.63 ± 0.22 | 0.49 ± 0.14 |
| | CNN | TVAE | 0.74 ± 0.12 | 0.50 ± 0.17 | 0.95 ± 0.10 | 0.67 ± 0.13 |
| | AdaBoost | Raw | 0.74 ± 0.10 | 0.53 ± 0.16 | 0.95 ± 0.14 | 0.69 ± 0.12 |
| | GNB | TVAE | 0.81 ± 0.11 | 0.72 ± 0.16 | 0.95 ± 0.10 | 0.80 ± 0.11 |
| | RF | SMOTE-NC | 0.80 ± 0.11 | 0.75 ± 0.15 | 0.89 ± 0.11 | 0.80 ± 0.11 |
| | MLP | CTGAN | 0.79 ± 0.11 | 0.78 ± 0.14 | 0.84 ± 0.16 | 0.80 ± 0.11 |
| | DT | CTGAN | 0.80 ± 0.11 | 0.75 ± 0.15 | 0.89 ± 0.11 | 0.80 ± 0.11 |
| | GNB | CopulaGAN | 0.83 ± 0.10 | 0.75 ± 0.15 | 0.95 ± 0.10 | 0.82 ± 0.11 |
| | Distil GPT2 (Table to Text) | Raw | 0.83 ± 0.10 | 0.88 ± 0.12 | 0.79 ± 0.21 | 0.83 ± 0.11 |
| | AdaBoost | CopulaGAN | 0.83 ± 0.10 | 0.88 ± 0.12 | 0.79 ± 0.21 | 0.83 ± 0.11 |
| | Transformer (Input Sequence) | SMOTE-NC | 0.88 ± 0.09 | 0.97 ± 0.07 | 0.68 ± 0.24 | 0.84 ± 0.11 |
| | GNB | SMOTE-NC | 0.84 ± 0.10 | 0.78 ± 0.14 | 0.95 ± 0.10 | 0.84 ± 0.11 |
| | T5-Small (Table to Text) | GPT4 | 0.85 ± 0.11 | 0.88 ± 0.08 | 0.84 ± 0.16 | 0.85 ± 0.11 |
| | Distil GPT2 (Input Sequence) | CopulaGAN | 0.86 ± 0.09 | 0.91 ± 0.09 | 0.79 ± 0.21 | 0.85 ± 0.10 |
| | BART-Base (Table to Text) | GPT4 | 0.91 ± 0.13 | 0.88 ± 0.11 | **1.00 ± 0.00** | 0.92 ± 0.09 |
| | RusBoost | CTGAN | 0.92 ± 0.07 | 0.94 ± 0.09 | 0.89 ± 0.11 | 0.92 ± 0.08 |
| | Distil GPT2 (Table to Text) | GPT4 | **0.93 ± 0.05** | 0.94 ± 0.04 | 0.95 ± 0.05 | **0.94 ± 0.06** |
| Heart Failure | BART-Base (Table to Text) | GPT4 | 0.09 ± 0.11 | 0.00 ± 0.00 | **1.00 ± 0.00** | 0.15 ± 0.11 |
| | MLP | TVAE | 0.53 ± 0.14 | 0.32 ± 0.16 | 0.78 ± 0.18 | 0.39 ± 0.13 |
| | FLAN-T5-Small (Table to Text) | GPT4 | 0.41 ± 0.12 | **1.00 ± 0.00** | 0.00 ± 0.00 | 0.45 ± 0.11 |
| | T5-Small (Table to Text) | GPT4 | 0.41 ± 0.11 | **1.00 ± 0.00** | 0.00 ± 0.00 | 0.45 ± 0.13 |
| | CNN | TVAE | 0.41 ± 0.14 | **1.00 ± 0.00** | 0.00 ± 0.00 | 0.45 ± 0.13 |
| | Transformer (Input Sequence) | SMOTE-NC | 0.41 ± 0.14 | **1.00 ± 0.00** | 0.00 ± 0.00 | 0.45 ± 0.13 |
| | RUSBoost | Raw | 0.51 ± 0.13 | 0.80 ± 0.11 | 0.22 ± 0.14 | 0.51 ± 0.12 |
| | ANN | TVAE | 0.53 ± 0.14 | 0.83 ± 0.13 | 0.22 ± 0.19 | 0.53 ± 0.14 |
| | Distil GPT2 (Table to Text) | Raw | 0.67 ± 0.13 | 0.98 ± 0.03 | 0.11 ± 0.14 | 0.54 ± 0.14 |
| | GNB | SMOTE-NC | 0.67 ± 0.13 | 0.56 ± 0.17 | **1.00 ± 0.00** | 0.61 ± 0.13 |
| | RF | TVAE | 0.68 ± 0.13 | 0.61 ± 0.17 | **1.00 ± 0.00** | 0.64 ± 0.13 |
| | DT | CTGAN | 0.71 ± 0.12 | 0.76 ± 0.15 | 0.89 ± 0.11 | 0.72 ± 0.12 |
| | DT | SMOTE-NC | 0.74 ± 0.12 | 0.85 ± 0.12 | 0.78 ± 0.18 | 0.77 ± 0.12 |
| | AdaBoost | TVAE | 0.82 ± 0.10 | 0.88 ± 0.11 | **1.00 ± 0.00** | 0.86 ± 0.10 |
| | DT | TVAE | 0.82 ± 0.10 | 0.88 ± 0.11 | **1.00 ± 0.00** | 0.86 ± 0.10 |
| | RusBoost | CopulaGAN | 0.82 ± 0.10 | 0.88 ± 0.11 | **1.00 ± 0.00** | 0.86 ± 0.10 |
| | Distil GPT2 (Table to Text) | GPT4 | **0.91 ± 0.04** | 0.95 ± 0.05 | **1.00 ± 0.00** | **0.94 ± 0.06** |

**Table 5:** Generalization assessment of the proposed framework on two additional datasets: Chronic Kidney Disease and Thyroid Cancer Recurrence. Performance is compared with machine learning, deep learning, and other language-model approaches under different data augmentation strategies. The best-performing model configurations are reported, with 95% confidence intervals for each evaluation metric. Ablation experiments further examine the contribution of individual framework components across these additional clinical prediction tasks.

| Dataset | Model | GAN | Precision | Specificity | Sensitivity | F1 Score |
| --- | --- | --- | --- | --- | --- | --- |
| Chronic Kidney Disease | CNN | TVAE | 0.14 ± 0.07 | 0.00 ± 0.00 | **1.00 ± 0.00** | 0.22 ± 0.10 |
| | FLAN-T5-Small (Table to Text) | GPT4 | 0.36 ± 0.11 | **1.00 ± 0.00** | 0.00 ± 0.00 | 0.42 ± 0.11 |
| | Transformer (Input Sequence) | TVAE | 0.36 ± 0.13 | **1.00 ± 0.00** | 0.00 ± 0.00 | 0.42 ± 0.13 |
| | RF | Raw | 0.66 ± 0.11 | 0.48 ± 0.12 | 0.89 ± 0.13 | 0.59 ± 0.14 |
| | MLP | CTGAN | 0.66 ± 0.13 | 0.48 ± 0.16 | 0.89 ± 0.11 | 0.59 ± 0.13 |
| | ANN | CTGAN | 0.72 ± 0.12 | 0.52 ± 0.16 | **1.00 ± 0.00** | 0.65 ± 0.12 |
| | Distil GPT2 (Table to Text) | Raw | 0.90 ± 0.08 | **1.00 ± 0.00** | 0.33 ± 0.11 | 0.69 ± 0.10 |
| | GNB | TVAE | 0.82 ± 0.10 | 0.78 ± 0.12 | **1.00 ± 0.00** | 0.83 ± 0.10 |
| | T5-Small (Table to Text) | GPT4 | 0.88 ± 0.12 | 0.91 ± 0.14 | 0.89 ± 0.14 | 0.89 ± 0.12 |
| | RusBoost | CTGAN | 0.98 ± 0.04 | **1.00 ± 0.00** | 0.89 ± 0.11 | 0.96 ± 0.08 |
| | BART-Base (Table to Text) | GPT4 | 0.98 ± 0.07 | **1.00 ± 0.00** | 0.89 ± 0.12 | 0.96 ± 0.12 |
| | RusBoost | TVAE | 0.98 ± 0.04 | **1.00 ± 0.00** | 0.89 ± 0.11 | 0.96 ± 0.08 |
| | AdaBoost | SMOTE-NC | 0.98 ± 0.04 | **1.00 ± 0.00** | 0.89 ± 0.11 | 0.96 ± 0.08 |
| | RF | SMOTE-NC | 0.98 ± 0.04 | **1.00 ± 0.00** | 0.89 ± 0.11 | 0.96 ± 0.08 |
| | DT | CTGAN | 0.98 ± 0.04 | **1.00 ± 0.00** | 0.89 ± 0.11 | 0.96 ± 0.08 |
|  | Distil GPT2 (Table to Text) | GPT4 | **1.00 ± 0.00** | **1.00 ± 0.00** | **1.00 ± 0.00** | **1.00 ± 0.00** |
| Thyroid Cancer Recurrence | Distil GPT2 (Table to Text) | Raw | 0.89 ± 0.07 | **1.00 ± 0.00** | 0.12 ± 0.08 | 0.55 ± 0.13 |
| | FLAN-T5-Small (Table to Text) | GPT4 | 0.69 ± 0.11 | 0.80 ± 0.08 | 0.65 ± 0.14 | 0.70 ± 0.13 |
| | ANN | TVAE | 0.74 ± 0.12 | 0.65 ± 0.16 | **1.00 ± 0.00** | 0.72 ± 0.12 |
| | T5-Small (Table to Text) | GPT4 | 0.86 ± 0.12 | 0.98 ± 0.09 | 0.41 ± 0.13 | 0.73 ± 0.14 |
| | AdaBoost | Raw | 0.75 ± 0.11 | 0.69 ± 0.12 | **1.00 ± 0.00** | 0.74 ± 0.16 |
| | MLP | CTGAN | 0.77 ± 0.12 | 0.80 ± 0.12 | 0.88 ± 0.11 | 0.78 ± 0.12 |
| | RF | SMOTE-NC | 0.93 ± 0.07 | 0.98 ± 0.03 | 0.76 ± 0.10 | 0.90 ± 0.08 |
| | DT | CTGAN | 0.92 ± 0.08 | 0.96 ± 0.04 | 0.88 ± 0.11 | 0.92 ± 0.08 |
| | BART-Base (Table to Text) | GPT4 | 0.95 ± 0.16 | 0.98 ± 0.12 | 0.88 ± 0.14 | 0.94 ± 0.12 |
| | CNN | SMOTE-NC | 0.95 ± 0.06 | 0.98 ± 0.03 | 0.88 ± 0.11 | 0.94 ± 0.07 |
| | RusBoost | TVAE | 0.95 ± 0.06 | 0.98 ± 0.03 | 0.88 ± 0.11 | 0.94 ± 0.07 |
| | Transformer (Input Sequence) | TVAE | 0.95 ± 0.06 | 0.98 ± 0.03 | 0.88 ± 0.11 | 0.94 ± 0.07 |
| | AdaBoost | CopulaGAN | 0.95 ± 0.06 | 0.98 ± 0.03 | 0.88 ± 0.11 | 0.94 ± 0.07 |
| | GNB | CopulaGAN | 0.95 ± 0.06 | 0.98 ± 0.03 | 0.88 ± 0.11 | 0.94 ± 0.07 |
| | Distil GPT2 (Table to Text) | GPT4 | **0.98 ± 0.04** | **1.00 ± 0.00** | 0.88 ± 0.04 | **0.96 ± 0.05** |

Across all four domains, traditional GAN-based augmentation methods such as CTGAN, TVAE, CopulaGAN, and SMOTE-NC improved performance for several baseline ML and neural network models; however, their results remained comparatively less stable and often demonstrated inconsistent sensitivity-specificity trade-offs. In contrast, the proposed context-aware table-to-text Distil GPT2 framework preserved clinical semantics while improving minority-class representation through controlled medical paraphrasing, resulting in more reliable and clinically consistent predictive behavior under highly limited and imbalanced data conditions. The ablation study suggests that Distil GPT2 achieved substantially improved performance with table-to-text conversion compared with raw table-input sequencing. Furthermore, incorporating paraphrasing to address class imbalance yielded the best overall performance, highlighting the effectiveness of combining table-to-text conversion with paraphrase-based augmentation. Additionally, the Distil GPT2-based Table-to-Text framework consistently demonstrated comparatively stable and narrower 95% CIs across multiple evaluation metrics and datasets, indicating lower uncertainty in the estimated performance under the evaluated test samples. For additional evaluation metrics of the proposed framework, including the area under the precision–recall curve (AUC-PR) and the area under the receiver operating characteristic curve (AUC-ROC), across the four datasets, please refer to **Table B.2** in the Supplementary section. For the performance of the other evaluated models on the four datasets, please refer to **Table B.3, Table B.4, Table B.5**, and **Table B.6**, respectively, in the Supplementary section. Additionally, **Table B.1** in the Supplementary section shows relatively stable performance of the proposed framework on the UCI Heart Disease dataset across varying numbers of positive training samples, suggesting no clear evidence of overfitting in the evaluated configurations.

### 4.2 Experimental Results: Contextual Understanding

**Table 6** compares the clinically important features emphasized by the proposed GPT-based framework with those that received minimal attribution from the highest-performing traditional ML and deep learning models across the four clinical domains. Despite the highly limited representation of positive and clinically important cases within the datasets, GPT-based modeling consistently identified medically meaningful variables strongly associated with disease progression and prediction outcomes. In contrast, several ML and neural network models assigned near-zero or substantially lower importance to multiple clinically relevant features, likely due to insufficient statistical representation of minority-class patterns in the training data.

**Table 6:** Comparison of clinically important features emphasized by GPT-based models and those underrepresented by the highest-performing traditional machine learning and deep learning approaches across different clinical domains. For DistilGPT-2, feature attributions were identified using gradient-based interpretability techniques, whereas for traditional machine learning and neural network models, explanations were derived using Shapley Additive Explanations (SHAP).

| Domain | Clinically Important Features Strongly Highlighted by GPT | Less Highlighted or Missed by ML/CNN Model | Clinical Significance (Doctors' Notes) |
| --- | --- | --- | --- |
| Heart Disease | Resting ECG Results, Exercise-Induced Angina, Resting Blood Pressure, Slope of ST Segment, Fasting Blood Sugar, Age | RUSBoost assigned near-zero or negligible importance to these variables | These features are well-established cardiovascular risk indicators associated with myocardial ischemia, hypertension-related vascular damage, metabolic dysfunction, and age-related coronary degeneration. GPT appears more aligned with established cardiology knowledge by emphasizing ischemic ECG abnormalities and exercise-related symptoms. |
| Heart Failure | Diabetes Mellitus, Anaemia, High Blood Pressure, Smoking Status | RUSBoost showed minimal marginal attribution to these features | These variables are clinically important contributors to heart failure progression through metabolic stress, reduced oxygen delivery, ventricular remodeling, and vascular injury. GPT better captured systemic comorbidities and lifestyle-related cardiovascular burden. |
| Chronic Kidney Disease | Hemoglobin, Specific Gravity, Blood Pressure, Serum Potassium, Age | AdaBoost assigned very low or near-zero importance to these biomarkers | These variables are central to chronic kidney disease pathology. Hemoglobin and RBC-related measures reflect renal anemia; specific gravity indicates tubular dysfunction; potassium imbalance reflects impaired renal excretion; hypertension accelerates nephron damage. GPT demonstrated stronger recognition of renal physiology and CKD progression markers. |
| Thyroid Cancer Recurrence | Tumor Focality, Smoking Status, Pathology Type, Thyroid Function Status, Radiotherapy History | CNN attributed minimal standalone importance to these factors | These features are clinically associated with recurrence risk, tumor aggressiveness, endocrine dysregulation, and treatment-related prognosis. GPT placed substantially greater emphasis on disease biology and clinical history, whereas CNN focused more narrowly on a limited subset of dominant predictive signals. |

Although predictive performance is essential for clinical prediction, high predictive scores alone do not necessarily indicate that a model captures clinically meaningful relationships or provides reliable risk assessment [5, 6]. In this study, contextual understanding is therefore considered complementary to predictive performance. Several models, including RUSBoost, AdaBoost, and CNN, achieved highly competitive predictive performance represented in **Table 4** and **Table 5**. However, their decisions still overlooked clinically meaningful relationships when positive cases are limited and the available data provide insufficient statistical representation presented in **Table 6**. Therefore, the proposed framework is evaluated not only based on predictive metrics but also on its ability to capture clinically meaningful patterns, with the subsequent reasoning and explainability analyses providing further insight into the clinical relevance of its predictions.

These findings suggest that conventional ML and neural network approaches primarily relied on dominant statistical correlations learned from limited samples, making it difficult to consistently capture weaker but clinically important relationships under severe class imbalance. Conversely, the Distil GPT2 table-to-text framework demonstrated stronger contextual understanding by recognizing clinically meaningful patterns through semantic and medical relevance rather than depending solely on direct statistical frequency. As a result, GPT-based modeling was better able to preserve and identify important disease-related signals even when positive-class representation was extremely limited, highlighting the advantage of contextual reasoning in low-resource clinical prediction settings.

## 5 DISCUSSION

This study suggests that converting structured clinical records into clinically meaningful text and leveraging GPT-based contextual reasoning can substantially improve disease prediction in low-resource and highly imbalanced healthcare datasets. Unlike conventional ML and neural network approaches that primarily depend on dominant statistical distributions, the proposed Distil GPT2 Table-to-Text framework preserved clinical semantics and identified medically meaningful relationships even when positive disease cases were severely underrepresented.

Across all four clinical domains, the proposed framework achieved more stable sensitivity, stronger generalization, and improved overall predictive balance compared with traditional ML, deep learning, transformer-based, and GAN-augmented approaches. The comparatively narrow CIs across several metrics further indicate improved robustness and reliability under limited-data conditions. Importantly, gradientbased interpretability analysis revealed that the GPT-based framework emphasized clinically relevant features associated with disease progression and risk, whereas several high-performing conventional models assigned minimal importance to important medical indicators despite achieving competitive statistical metrics.

These findings suggest that purely correlation-driven learning may not always be sufficient for reliable clinical prediction under severe class imbalance. By integrating contextual medical reasoning with explainable prediction mechanisms, the proposed framework moves beyond blackbox classification toward clinically interpretable and reliable decision support. This capability is particularly important for real-world healthcare systems, where early identification of high-risk patients must be accompanied by transparent reasoning and clinically meaningful evidence to support physician trust and safe deployment. Although feature importance should not be interpreted solely as evidence of causal relationships, it can help identify potential risk factors associated with a condition or disease. These factors can support early risk prediction and encourage individuals to seek timely medical screening. Therefore, feature-importance analysis could contribute to the development of cost-effective, large-scale prevention strategies.

The proposed framework is intended primarily as a clinical decision-support tool rather than a replacement for physician judgment. By providing patient-level risk predictions along with clinically meaningful feature-level explanations, the framework could assist clinicians in identifying higher-risk patients, prioritizing individuals for further assessment, and supporting earlier preventive or therapeutic interventions. This can be particularly relevant in low-resource settings where access to specialist expertise and advanced clinical decision-support systems is limited. However, the present study evaluates predictive performance using retrospective public datasets and does not directly assess its impact on physician decision-making, clinical workflow, or patient outcomes. Future work should therefore focus on prospective clinical evaluation to determine whether model-assisted decision-making can improve physician risk assessment, reduce decision-making errors, and support more consistent clinical decisions. Several challenges should also be considered before real-world deployment, including differences in patient populations and clinical settings, changes in data distributions over time, computational requirements, data privacy, Electronic Health Record integration, and model reliability. Regular monitoring and updating will be necessary to maintain performance, while appropriate clinician oversight will be important to avoid over-reliance on automated predictions. Therefore, external and prospective validation in real-world clinical environments will be necessary before routine clinical use.

Although newer and more capable LLMs are now available, we selected Distil GPT2 because it is a compact 82-million-parameter transformer that provides a lightweight and computationally efficient alternative to larger language models [37]. This choice allowed us to evaluate the proposed table-to-text framework under relatively modest computational requirements, rather than to claim that Distil GPT2 represents the current stateof-the-art LLM. Newer lightweight and quantized models provide promising alternatives for future investigation, particularly for improving scalability and deployment efficiency.

Overall, this study highlights the potential of GPT-based table-to-text modeling as a practical and clinically interpretable framework for early disease-risk screening using limited structured healthcare data, while recognizing the need for external and prospective clinical validation before real-world deployment.

## 6 CONCLUSION

This study suggests that GPT-based table-to-text modeling can provide a practical and clinically reliable approach for disease prediction from small and highly imbalanced structured healthcare datasets. By transforming tabular clinical records into medically meaningful text and integrating controlled clinical paraphrasing with explainable contextual reasoning, the proposed framework achieved improved sensitivity, stable generalization, and more clinically relevant feature identification compared with conventional ML and deep learning approaches. Importantly, the model not only improved predictive performance but also provided transparent reasoning through clinically meaningful indicators, supporting physician trust and interpretability in medical decision-making. These findings highlight the realworld potential of context-aware language models for early risk detection, reliable clinical decision support, and deployment in low-resource healthcare settings where limited data availability often restricts the effectiveness of traditional predictive systems. In conclusion, when clinical data have limited representation of relationships between input features and disease outcomes, the proposed Distil GPT2–based table-to-text approach, particularly when combined with paraphrasing, can effectively capture clinically relevant relationships compared with conventional ML and neural network models. Although Distil GPT2–based modeling has higher computational requirements than conventional approaches, its computational cost remains manageable for small, low-volume clinical datasets, making it a potentially practical option for such settings.

## Data Availability

UC Irvine Machine Learning Repository

https://archive.ics.uci.edu/

## 7 DECLARATIONS

### 7.1 Ethics Approval

This study involved secondary analysis of four publicly available, de-identified clinical datasets: Heart Disease, Heart Failure Clinical Records, Chronic Kidney Disease, and Differentiated Thyroid Cancer Recurrence. No new data were collected from human participants, and no identifiable personal information was accessed. Ethical approval was not required for this secondary analysis under the applicable institutional requirements.

### 7.2 Consent to Participate

Not applicable. This study used publicly available, de-identified datasets and did not involve direct recruitment of participants.

### 7.3 Consent to Publish

Not applicable. This manuscript does not contain identifiable information about individual participants.

## 7.6 Data Availability

The four clinical datasets analyzed in this study are publicly available through the UCI Machine Learning Repository:

- **Heart Disease:** https://archive.ics.uci.edu/dataset/45/heart+disease
- **Heart Failure Clinical Records:**https://archive.ics.uci.edu/dataset/519/heart+failure+clinical+records
- **Chronic Kidney Disease:** https://archive.ics.uci.edu/dataset/336/chronic+kidney+disease
- **Differentiated Thyroid Cancer Recur-rence:** https://archive.ics.uci.edu/dataset/915/differentiated+thyroid+cancer+recurrence

The datasets were used in accordance with their respective access, licensing, and data-use conditions.

