## Supplementary for "Can GPT Be Used as an Alternative to Traditional Machine Learning and Neural Networks for Prediction on Small Clinical Datasets?"

### Supplementary A. ADDITIONAL EXAMPLES OF CLINICAL TEXT CONVERSION AND GENERATION

This supplementary section provides additional examples of converting structured clinical data into natural-language text and generating synthetic positive-class samples through medically relevant paraphrasing. The examples demonstrate how the proposed approach preserves the original clinical information, disease labels, and medical context while producing diverse text representations for the four datasets.

#### Supplementary A.1. Example from Heart Disease

| Age (years) | Sex (1 = Male, 0 = Female) | Chest Pain Type (1 = Typical Angina, 2 = Atypical Angina, 3 = Non-anginal Pain, 4 = Asymptomatic) | Resting Blood Pressure (mm Hg) | Serum Cholesterol (mg/dL) | Fasting Blood Sugar 120 mg/dL, 1 = True, 0 = False) | Resting ECG Results (0 = Normal, 1 = ST-T Wave Abnormality, 2 = Left Ventricular Hypertrophy) | Maximum Heart Rate Achieved | Exercise-Induced Angina (1 = Yes, 0 = No) | ST Depression Induced by Exercise (mm) | Slope of Peak Exercise ST Segment (1 = Upsloping, 2 = Flat, 3 = Downsloping) | Number of Major Vessels (0 = 3) Colored by Fluoroscopy | Thalassemia (3 = Normal, 6 = Fixed Defect, 7 = Reversible Defect) | Heart Disease (0 = No Disease, 1 = Heart Disease) |
|---|---|---|---|---|---|---|---|---|---|---|---|---|---|
| 65-69 | 1 | 4 | 160 | 286 | 0 | 2 | 108 | 1 | 1.5 | 2 | 3.0 | 3.0 | 1 |

**Table to Text:**

Patient is a male aged 65-69 years, presenting with chest pain type 4, resting blood pressure 160 mm Hg, serum cholesterol 286 mg/dL, normal fasting blood sugar, resting ECG showing result Left Ventricular Hypertrophy, maximum heart rate achieved 108, with exercise-induced angina, ST depression of 1.5 mm, Slope of Peak Exercise ST Segment: Flat, 3.0 major vessels affected, thalassemia type Normal.

**Synthetic Data:**

A 65-69-year-old male with chest pain type 4 presents with normal fasting blood sugar. Resting blood pressure is 160 mm Hg, and serum cholesterol is 286 mg/dL. Maximum heart rate achieved is 108. ST depression is 1.5 mm, with a flat ST segment. ECG shows Left Ventricular Hypertrophy, and there is exercise-induced angina. Three major vessels are affected, and thalassemia is of the Normal type.

**Figure A.1:** An example of converting tabular data into text and generating synthetic positive-class samples for the Heart Disease dataset through text paraphrasing while maintaining medical relevance and class labels.

**Supplementary A.2. Example from Heart Failure**

| age (years) | anemia (0 = No, 1 = Yes) | creatinine_ phosphokinase (IU/L) | diabetes (0 = No, 1 = Yes) | ejection_ fraction (%) | high_blood_ pressure (0 = No, 1 = Yes) | platelets (per µL) | serum_ creatinine (mg/dL) | serum_ sodium (mEq/L) | sex (1 = Male, 0 = Female) | smoking (0 = No, 1 = Yes) | time (days) | death_event (1 = Death event, 0 = No death event) |
|---|---|---|---|---|---|---|---|---|---|---|---|---|
| 75–79 | 0 | 582 | 0 | 20 | 1 | 265000.0 | 1.9 | 130 | 1 | 0 | 4 | 1 |

### Table to Text:

Patient is a 75–79-year-old male with an ejection fraction of 20%, serum creatinine level of 1.9 mg/dL, serum sodium level of 130 mEq/L, platelets count of 265000.0 per µL, creatinine phosphokinase level of 582 IU/L, and has high blood pressure. The patient is not diabetic, does not have anaemia, and does not smoke. The follow-up duration is 4 days.

### Synthetic Data:

The male patient, aged 75–79, has a serum creatinine of 1.9 mg/dL and a sodium level of 130 mEq/L. Their ejection fraction is 20%, and creatinine phosphokinase is 582 IU/L. Platelets count is 265,000 per µL. They are hypertensive, not diabetic, have no anemia, and do not smoke. Follow-up duration is 4 days.

**Figure A.2:** An example illustrating the conversion of tabular Heart Failure data into clinical text and the generation of synthetic positive-class samples through clinical text paraphrasing while preserving medical relevance and the original class labels.

#### Supplementary A.3. Example from Chronic Kidney Disease

| Age (years) | Blood Pressure (mm Hg) | Specific Gravity of Urine | Albumin (0 - None, 1 - Trace, 2 - Mild, 3 - Moderate, 4 - Severe, 5 - Very Severe) | Sugar (0 - None, 1 - Mild, 2 - Moderate, 3 - High, 4 - Very High) | Red Blood Cells (1 - Normal, 2 - Mildly Abnormal, 3 - Abnormal) | Pus Cells (0 - Abnormal, 1 - Normal) | Pus Cell Clumps (0 - Not 1 - Present) | Bacteria (0 - Not Present, 1 - Present) | Blood Glucose Random (mg/dL) | Urea (mg/dL) | Serum Creatinine (mg/dL) | Sodium (mEq/L) | Potassium (mEq/L) | Hemoglobin (g/dL) | Packed Cell Volume (%) | White Blood Cell Count (cells/cumm) | Red Blood Cell Count (million/ cumm) | Hypertension (1 - Yes, 0 - No) | Diabetes Mellitus (1 - Yes, 0 - No) | Coronary Artery Disease (1 - Yes, 0 - No) | Appetite (1 - Poor, 0 - Good) | Pedal Edema (1 - Yes, 0 - No) | Anemia (1 - Yes, 0 - No) | Chronic Kidney Disease (1 - Yes, 0 - No) |
|---|---|---|---|---|---|---|---|---|---|---|---|---|---|---|---|---|---|---|---|---|---|---|---|---|
| 45–49 | 70.0 | 1.005 | 4.0 | 0.0 | 1 | 0 | 1 | 0 | 117.0 | 56.0 | 3.8 | 111.0 | 2.5 | 11.2 | 32.0 | 6700.0 | 3.9 | 1 | 0 | 0 | 1 | 1 | 1 | 1 |

### Table to Text:

Patient aged 45–49 years with blood pressure of 70.0 mm Hg, urine specific gravity of 1.005, severe albumin level and no urine sugar. Red blood cells are normal, pus cells are abnormal, pus cell clumps are present, and bacteria are absent. Random blood glucose is 117.0 mg/dL, urea is 56.0 mg/dL, serum creatinine is 3.8 mg/dL, sodium is 111.0 mEq/L, and potassium is 2.5 mEq/L. Hemoglobin is 11.2 g/dL, packed cell volume is 32.0%, white blood cell count is 6700.0 cells/cumm, and red blood cell count is 3.9 million/cumm. The patient has hypertension, poor appetite, pedal edema, and anemia, but no diabetes mellitus or coronary artery disease.

### Synthetic Data:

A patient aged 45–49 years has hypertension, poor appetite, pedal edema, and anemia, with no diabetes mellitus or coronary artery disease. Blood pressure is 70.0 mm Hg. Urine findings include specific gravity of 1.005, severe albumin level, no sugar, normal red blood cells, abnormal pus cells, present pus cell clumps, and absent bacteria. Random blood glucose is 117.0 mg/dL and urea is 56.0 mg/dL. Serum creatinine is 3.8 mg/dL, sodium is 111.0 mEq/L, and potassium is 2.5 mEq/L. Hemoglobin is 11.2 g/dL, packed cell volume is 32.0%, white blood cell count is 6700.0 cells/cumm, and red blood cell count is 3.9 million/cumm.

**Figure A.3:** Illustration of the table-to-text transformation and minority-class augmentation process for the Chronic Kidney Disease dataset, where clinically consistent paraphrases are generated for positive cases while retaining their underlying medical information and class assignments.

**Supplementary A.4. Example from Thyroid Cancer Recurrence**

| Age (years) | Gender (0 = Female, 1 = Male) | Smoking (No, 1 - Yes) | History of Radiotherapy (No, 1 - Yes) | History of Chemotherapy (0 - No, 1 - Yes) | Thyroid Function (0 = Euthyroid, 1 = Subclinical Hyperthyroidism, 2 = Subclinical Hypothyroidism, 3 = Clinical Hypothyroidism) | Physical Examination (0 = Normal, 1 - Single nodule, 2 - Single + multiple nodules, 3 - Multiple nodules, 4 - Extensive, 5 - Multinodular goiter, 6 = Diffuse goiter) | Adenopathy (0 = No, 1 - Left, 2 - Right, 3 - Bilateral, 4 - Extensive, 5 - Posterior) | Pathology Type (0 = Papillary, 1 - Micropapillary, 2 = Follicular, 3 = Hurthle cell) | Tumor Focality (0 = Uni-focal, 1 = Multi-focal) | AJCC Size / Extent (0 = Tis, 1 = T1a, 2 = T1b, 3 = T2, 4 = T3a, 5 = T3b, 6 = T4a, 7 = T4b) | Lymph Node Involvement (N stage: 0 = No, 1 = N1a, 2 = N1b) | Metastasis (M stage: 0 = No, 1 = M0, 2 = M1) | AJCC Stage (0 = I, 1 = Stage II, 2 = Stage III, 3 = Stage IV) | Response to Initial Therapy (0 = Excellent, 1 = Indeterminate, 2 = Biochemical Incomplete, 3 = Structural Incomplete) | Disease Recurrence (0 = No, 1 = Yes) |
|---|---|---|---|---|---|---|---|---|---|---|---|---|---|---|---|
| 25–29 | 0 | 0 | 0 | 0 | 0 | 0 | 4 | 1 | 1 | 1 | 0 | 2 | 0 | 1 | 1 |

### Table to Text:

Patient is a 25–29-year-old female with no history of smoking, radiotherapy, or chemotherapy. Thyroid function is euthyroid, and physical examination is normal. Adenopathy is extensive. Pathology is micropapillary, and the tumor is multifocal. Tumor stage is T1a, with no lymph node involvement and an M1 metastasis classification. The overall AJCC stage is I. Response to initial therapy is indeterminate.

### Synthetic Data:

A female patient aged 25–29 years has no history of smoking, radiotherapy, or chemotherapy. She has euthyroid thyroid function and normal physical examination findings, with extensive adenopathy. The pathology is micropapillary, and tumor focality is multifocal. The tumor is classified as T1a, with no lymph node involvement and metastasis classified as M1. Overall AJCC stage is I, and the response to initial therapy is indeterminate.

**Figure A.4:** Representative example from the Thyroid Cancer Recurrence dataset demonstrating how structured patient records is expressed as descriptive clinical narratives and how minority recurrence cases are expanded using medically consistent paraphrases without changing their clinical meaning or assigned outcomes.

### Supplementary B. ADDITIONAL EXPERIMENTAL RESULTS

This supplementary section provides supplementary results for the four clinical datasets, including the best-performing configurations of the evaluated machine learning and neural network models and additional precision–recall curve (AUC-PR) and area under the receiver operating characteristic curve (AUC-ROC) results for the proposed framework. It also presents results for the UCI Heart Disease dataset with varying numbers of positive training samples to examine performance stability across different train–test configurations.

#### Supplementary B.1. Different Train/Test Formulations

**Table B.1:** Performance of the proposed framework on the UCI Heart Disease dataset with varying numbers of positive training samples. The model was trained separately for each ratio, with strict separation of training and test data to prevent data leakage.

| **Dataset** | **Variation** | **Train negative** | **Train positive** | **Test negative** | **Test positive** | **Train : Test ratio** | **Negative Recall (%)** | **Heart Disease Recall (%)** |
|---|---|---|---|---|---|---|---|---|
| Heart Disease | 10 positive: train → test | 128 | 108 | 32 | 29 | 79:21 | 94 | 100 |
| Heart Disease | 20 positive: train → test | 128 | 98 | 32 | 39 | 76:24 | 91 | 90 |
| Heart Disease | **Original** | 128 | 118 | 32 | 19 | 83:17 | 94 | 95 |
| Heart Disease | 10 positive: test → train | 128 | 128 | 32 | 9 | 86:14 | 97 | 89 |

#### Supplementary B.2. Additional Performance Metrics Across the Four Clinical Datasets

**Table B.2:** Area under the precision–recall curve (AUC-PR) and area under the receiver operating characteristic curve (AUC-ROC) of the proposed framework across the four clinical datasets.

| **Dataset** | **Model** | **Sampling** | **AUC-PR (%)** | **AUC-ROC (%)** |
|---|---|---|---|---|
| Heart Disease | Distil GPT2 (Table to Text) | GPT4 | 98 | 99 |
| Heart Failure Clinical Records | Distil GPT2 (Table to Text) | GPT4 | 95 | 98 |
| Chronic Kidney Disease | Distil GPT2 (Table to Text) | GPT4 | 100 | 100 |
| Differentiated Thyroid Cancer Recurrence | Distil GPT2 (Table to Text) | GPT4 | 96 | 99 |

#### Supplementary B.3. Results from Heart Disease

**Table B.3:** Best-performing configurations of the evaluated machine learning and neural network models across GAN-based augmentation methods for the UCI Heart Disease dataset.

| **Model** | **Sampling Technique** | **Negative Recall (%)** | **Heart Disease Recall (%)** | **Balanced Accuracy (%)** |
|---|---|---|---|---|
| ANN | SMOTE-NC | 41 | 63 | 52.0 |
| CNN | TVAE | 50 | **95** | 72.5 |
| MLP | CTGAN | 78 | 84 | 81.0 |
| RF | SMOTE-NC | 75 | 89 | 82.0 |
| DT | CopulaGAN \| CTGAN | 69 \| 75 | **95** \| 89 | 82.0 |
| AdaBoost | SMOTE-NC \| CopulaGAN | **88** | 79 | 83.5 |

| **Model** | **Sampling Technique** | **Negative Recall (%)** | **Heart Disease Recall (%)** | **Balanced Accuracy (%)** |
|---|---|---|---|---|
| RUSBoost | CopulaGAN | **88** | 79 | 83.5 |
| GNB | CopulaGAN | 75 | **95** | **85.0** |

**Supplementary B.4. Results from Heart Failure**

**Table B.4:** Performance of the best configuration of each evaluated machine learning and neural network model using GAN-based data augmentation on the Heart Failure Clinical Records dataset.

| **Model** | **Sampling Technique** | **Negative Recall (%)** | **Positive Recall (%)** | **Balanced Accuracy (%)** |
|---|---|---|---|---|
| CNN | SMOTE-NC \| CTGAN \| CopulaGAN \| TVAE | **100** \| 0 \| 0 \| **100** | 0 \| **100** \| **100** \| 0 | 50.0 |
| ANN | TVAE | 83 | 22 | 52.5 |
| MLP | CopulaGAN \| TVAE | 10 \| 32 | **100** \| 78 | 55.0 |
| GNB | SMOTE-NC | 56 | **100** | 78.0 |
| RF | TVAE | 61 | **100** | 80.5 |
| DT | TVAE | 88 | **100** | **94.0** |
| AdaBoost | TVAE | 88 | **100** | **94.0** |
| RUSBoost | CopulaGAN | 88 | **100** | **94.0** |

**Supplementary B.5. Results from Chronic Kidney Disease**

**Table B.5:** Performance comparison of machine learning and neural network models under their best-performing GAN-based augmentation configurations on the Chronic Kidney Disease dataset.

| **Model** | **Sampling Technique** | **Negative Recall (%)** | **Kidney Disease Recall (%)** | **Balanced Accuracy (%)** |
|---|---|---|---|---|
| CNN | All four techniques | **100** \| 0 | 0 \| **100** | 50.0 |
| MLP | CTGAN | 48 | 89 | 68.5 |
| ANN | CTGAN | 52 | **100** | 76.0 |
| GNB | TVAE | 78 | **100** | 89.0 |
| RF | CopulaGAN / SMOTE-NC | **100** | 89 | **94.5** |
| DT | All four techniques | **100** | 89 | **94.5** |
| AdaBoost | All four techniques | **100** | 89 | **94.5** |
| RUSBoost | All four techniques | **100** | 89 | **94.5** |

**Supplementary B.6. Results from Thyroid Cancer Recurrence**

**Table B.6:** Comparison of the best-performing configurations of machine learning and neural network models evaluated with GAN-based data augmentation on the Differentiated Thyroid Cancer Recurrence dataset.

| **Model** | **Sampling Technique** | **Negative Recall (%)** | **Cancer Recurrence Recall (%)** | **Balanced Accuracy (%)** |
|---|---|---|---|---|
| ANN | TVAE | 65 | **100** | 82.5 |
| MLP | CopulaGAN \| CTGAN | 80 | 88 | 84.0 |
| RF | SMOTE-NC | **98** | 76 | 87.0 |
| DT | CopulaGAN \| CTGAN | 96 | 88 | 92.0 |
| CNN | SMOTE-NC | **98** | 88 | **93.0** |

| **Model** | **Sampling Technique** | **Negative Recall (%)** | **Cancer Recurrence Recall (%)** | **Balanced Accuracy (%)** |
|---|---|---|---|---|
| GNB | CTGAN \| CopulaGAN | **98** | 88 | **93.0** |
| AdaBoost | CTGAN \| CopulaGAN | **98** | 88 | **93.0** |
| RUSBoost | TVAE \| CopulaGAN \| CTGAN | **98** | 88 | **93.0** |
